# Modelling long-term COVID-19 hospital admission dynamics using empirical immune protection waning data

**DOI:** 10.1101/2022.05.16.22275130

**Authors:** Bastien Reyné, Mircea T. Sofonea, Samuel Alizon

## Abstract

Immune waning is key to the timely anticipation of COVID-19 long-term dynamics. We assess the impact of periodic vaccination campaigns using a compartmental epidemiological model with embedded multiple age structures and empiric time-dependent vaccine protection kinetics. Despite the uncertainty inherent to such scenarios, we show that vaccination campaigns decreases the yearly number of COVID-19 admissions. However, especially if restricted to individuals over 60 years old, vaccination on its own seems insufficient to prevent thousands of hospital admissions and it suffers the comparison with non-pharmaceutical interventions aimed at decreasing infection transmission. The combination of such interventions and vaccination campaigns appear to provide the greatest reduction in hospital admissions.

## 1 Introduction

From the beginning, COVID-19 pandemic management had to deal with numerous unknowns and strongly relied on mathematical modelling to guide non-pharmaceutical interventions (NPIs) implementation [Kucharski et alii, 2020]. The first steps of the COVID-19 management due to the SARS-CoV-2 emergence led to exploring some possibilities. For instance, some considered the possibility to reach herd immunity — thus terminating the epidemic provided a sufficient proportion of the population is immune to the disease — as a perennial replacement of the emergency use of lock-downs and other strong non-pharmaceutical interventions (NPIs) to alleviate the hospital burden. These considerations happened at a time when vaccines were not available. Given the estimates of the proportion of the population needed to be immune to reach the said threshold, it would have required a massive infection of the population [Kwok et alii, 2020; Randolph and Barreiro, 2020; Fontanet and Cauchemez, 2020], and hence a non-negligible death toll in different populations.

In late 2020, the availability of safe and efficient vaccines provided a powerful tool for pandemic management. Furthermore, it was realised that in addition to protecting against severe symptoms, vaccination also protected against infection itself and even reduced transmission rate leading to the hope that we could eradicate, or at least easily control, the virus. However, the hope was a little dampened by the emergence of the Alpha variant of concern (VOC) that was both more transmissible and more virulent than the ancestral lineages [Davies, Abbott, et alii, 2021; Davies, Jarvis, et alii, 2021]. And it also reminded virus evolution cannot be taken out of the equation [Alizon and Sofonea, 2021]. The emergence of the Delta VOC a few months later — once again both more transmissible and more virulent [Fisman and Tuite, 2021; Alizon, Haim-Boukobza, et alii, 2021] — combined with the vaccine efficacy *ab initio* cast some doubt on the strategy to be adopted. Later on, the overall SARS-CoV-2 reported time-induced loss of immunity entombed the possibility for herd immunity.

In addition to this emergence of variant issue, field data rapidly showed that immunity against SARS-CoV-2 wanes over time. For instance, protection against Omicron VOC severe forms decreases from ∼90% to ∼73% in six months for individuals with a full vaccination schedule (2 doses with Pfizer for 65+ y.o. individuals) [UKHSA, 2022]. For the protection against infection, the decrease is even more pronounced [UKHSA, 2022]. This is consistent with numerous studies on antibody titres that show a decline with time [Peng et alii, 2022; Pérez-Alós et alii, 2022; Levin et alii, 2021]. This problem is exacerbated by the fact that some VOCs are particularly prone to evading pre-existing immune responses [Dejnirattisai et alii, 2022]. Note that individual histories become particularly important to anticipate the efficiency of the immune response [Reynolds et alii, 2022].

In France, the current management strategy consists in vaccinating the population as much as possible and implementing NPIs when epidemic waves when hospital capacity approaches its limits. Mathematical modelling has provided in many countries and all along the pandemic useful insights and good approximations to assess what could happen in the near future if the current conditions remain the same. For instance, the first waves and the emergence of the Alpha, Delta and Omicron VOCs were quite rapidly followed by modelling studies providing useful insights to guide public policies [Salje et alii, 2020; Domenico et alii, 2021; Alizon, Haim-Boukobza, et alii, 2021; Sofonea, Roquebert, et alii, 2022]. However, past one month, the projections made become less precise and past few months some mathematical and biological assumptions might rapidly alter the general trends [Sofonea and Alizon, 2021]. That being said, we believe such long-term trends could still be of interest to the authorities that would consider the possibility of long-term management. Another proof-of-concept point would be to correct some limitations of many mathematical models based on the usual set of ordinary differential equations (ODEs) [Sofonea, Reyné, et alii, 2021].

Thanks to the work and dedication of some people, e.g. at UKHSA, this pandemic generates data that is unique in the history of epidemics, such as the vaccine efficacy over time since vaccination for the different variants circulating in the population. Such data can help us anticipate long-term trends. However, usual ODEs-based models are inadequate to integrate empirical data that depart too much from the exponential distribution, which is the case for immune protection waning.

Because of these technical difficulties, long-term projections are scarce in the literature. In the French context, Bosetti et alii [2022], studied under what booster administration and NPIs implementation a new epidemic could be contained. However, their work was done before the Omicron VOC, and only on a six-month horizon. Kissler et alii [2020] investigated the potential long-term effects of the seasonality observed in other epidemics than that of SARS-CoV-2. They highlighted the need for NPIs, but their results were obtained before vaccine implementation and the emergence of VOCs. Saad-Roy et alii [2021] studied different vaccine administration patterns for several scenarios of immunity duration. However, their long-term insights were very uncertain because, at the time, there was no data on immune protection waning. Finally, Ghosh et alii [2022] used a non-Markovian setting to capture immunity waning but their scenarios also remained on a few months’ horizon.

In this work, by extending a non-Markovian approach [Reyné et alii, 2022] that readily accounts for the time spent in each compartment, we leverage the field data on immune protection from UKHSA and qualitatively explore long-term epidemic dynamics. We focus on the case of France but it reflects that of many high-income countries in terms of vaccination coverage and age distributions. In our scenarios, we account for immunity waning as well as Omicron-specific phenotypic traits and compare three vaccination campaign strategies. Furthermore, we also investigate the impact of non-pharmaceutical interventions (NPIs) at the whole population level, such as mask-wearing or air quality improvement, that can decrease the transmission rate of the infection.

## 2 Methods

### An epidemiological model with time structures

We build on an existing non-Markovian epidemiological compartmental model that accounts for time since entry in the different compartments [Reyné et alii, 2022]. The main extension is that we implement vaccine booster doses and loss of immunity over time for vaccinated, boosted, or recovered individuals (Figure 1A). Susceptible individuals of age *a* (the density of which is denoted *S*_*a*_) can either become fully vaccinated (*V*_*a*_), or contract a mild 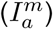 or severe infection 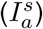 Mildly-infected individuals always recover and move to the compartment *R*_*a*_. Both recovered and vaccinated individuals can be (re)infected, but at a reduced rate compared to susceptible individuals (see also the section below about parameterisation for further details about the biological assumptions made). If this (re)infection is mild, individuals move to a separate compartment 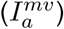 to account for a potential immunity-induced reduction in infectiousness. Vaccinated and recovered individuals may be (re)vaccinated and move to the booster compartment (*B*_*a*_), where their protection increases. Finally, severely infected 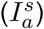 and previously immunised mildly infected individuals 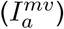 also end up in the booster compartment upon recovery, assuming high protection against potential new infection. Overall, the boosted compartment consists of individuals with booster vaccination dose(s), two natural infections, one vaccination and one infection, or having recovered from a severe infection.

**Figure 1:**
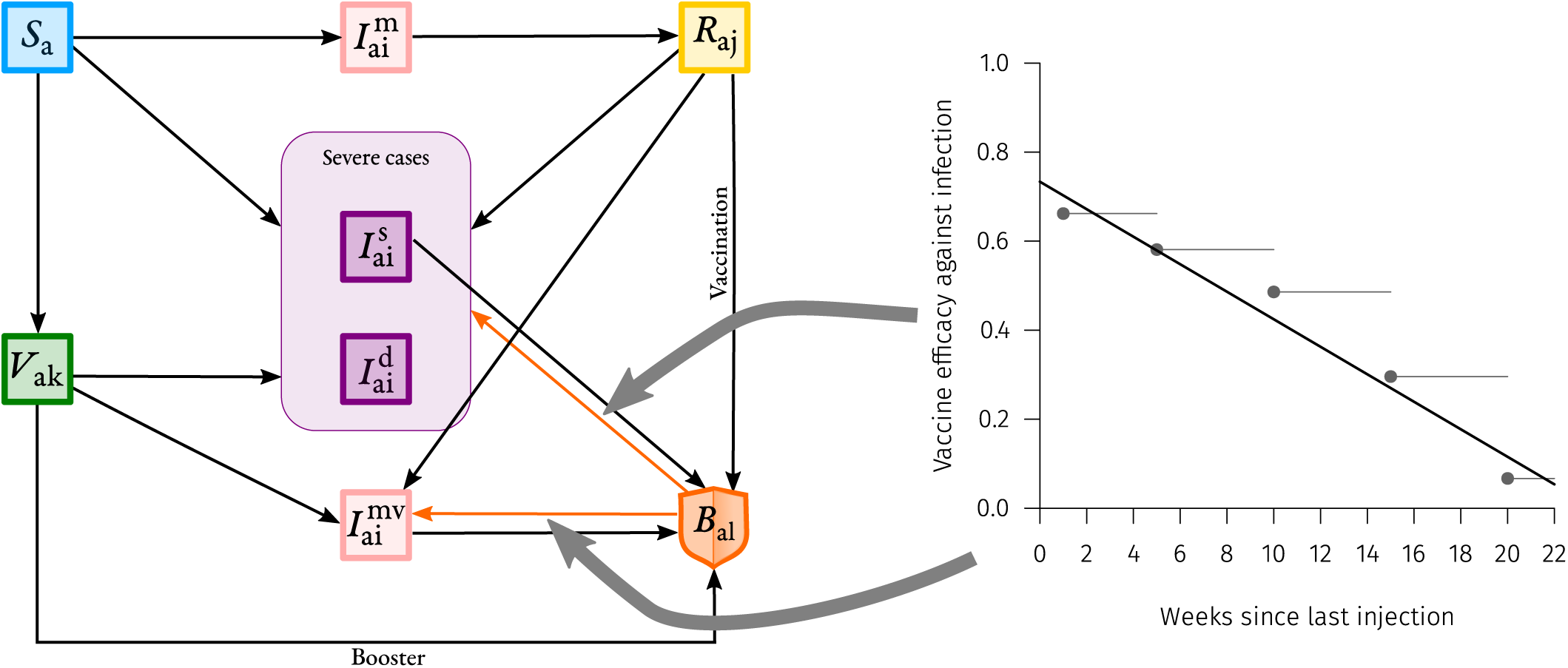
Description of the memory-based model with immune waning. **A)** Flowchart of the compartmental model. Arrows show transitions between compartments. *S*_*a*_ stands for Susceptible individuals of age 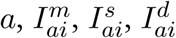 and 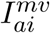 stand respectively for mildly/severely/severely-that-will-die/mildly-partly-immune infected individuals of age *a* infected since *i* days, *V*_*ak*_ stands for Vaccinated individuals of age *a* vaccinated *k* days ago, *R*_*aj*_ stands for Recovered individuals of age *a* that cleared the disease *j* days ago and finally *B*_*aℓ*_ stands for individuals of age *a* that received a booster vaccine dose *ℓ* days ago. Orange arrows show some of the transitions that depend on the time spent in the compartment (here *ℓ* days) that are parametrized through real immune waning data shown on panel B. **B)** Waning of vaccine efficacy against infection. Dots corresponds to real data from UKHSA [2022] for the Pfizer/BioNTech vaccine (BNT162b2) after a booster dose. The lines correspond to the baseline of the immunity decrease model implementation and the shaded areas to the uncertainty used within the sensitivity analysis. Here, we show the protection against an Omicron/BA.1 VOC infection for individuals that received a booster dose. Large grey arrows indicate where the waning data shown acts in the compartmental model.

The model accounts for memory effects, meaning that we record the time spent by the individuals in each compartment. Knowing the time since vaccination (*k*) for vaccinated individuals, the time since clearance (*j*) for recovered individuals, and the time since the entry into the booster compartment (*ℓ*), allows us to readily account for the waning in immune protection. This particularity is modelled using partial differential equations, that provide for each compartment the rate at which individuals enter the compartment with a boundary equation and the time-dependent depart of the compartment through the differential equation. For instance, for the vaccinated compartment we have the boundary equation

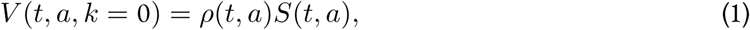

coupled with the differential equation

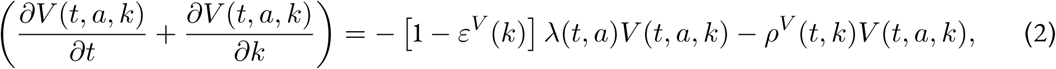

where *ρ*(*t, a*) is the initial vaccination rate (1^st^ and 2^nd^ doses), *ε*^*V*^ (*k*) is the vaccine efficacy *k* days after the first dose, *λ*(*t, a*) is the force of infection and *ρ*^*V*^ (*k*) corresponds to the third dose vaccination rate. The complete set of equations is available in Appendix S2.

### Fitting procedure and transmission rates

To simulate long-term scenarios, we need to generate a population with a heterogeneous exposition background to SARS-CoV-2 infection and vaccination. This is done by fitting the dynamics of the daily COVID-19 hospital admissions, which correspond to the severe cases in our model. The fit procedure is based on ordinary least squares and is done piecewise (the length of pieces varies, depending on important changes in transmission rates).

Transmission rates were fitted from the beginning of the simulations (Jan 1, 2021) up to May 6, 2022. The last fitted value corresponds to the baseline transmission rate at the end of the simulations. This is not completely neutral since the fitting account for NPIs or spontaneous behavioural changes in the reflection of the epidemic. In France, at that time the restrictions were quite low, only mask-wearing in public transportations was required so it reflects a situation with very scarce epidemic control amongst the population.

External factors, such as the weather, are known to impact transmission dynamics up to ~17% [Ma et alii, 2021]. We decided to include a seasonality by assuming sinusoidal variations such that in summer the transmission rate is decreased to –10% and increased by +10% in the winter. Since this seasonality is largely driven by behavioural changes, we assume that it applies similarly to all variants.

Finally, in some scenarios, we further decrease the baseline transmission rate by 20%. This is done to assess both the potential impact of long-term NPIs (such as improving indoor air quality) and to test the sensitivity of the model regarding this parameter.

### Virus related parameters

The presented model is not multi-strain, each time a new VOC becomes more prevalent it provokes a switch in the model parameters. We also neglect virus evolution meaning the current resident strain (Omicron BA.2 VOC) will remain till the end of simulations. The virus-related parameters are available in Table 1.

**Table 1:** Description of the Omicron-related model parameters and their default value. For each parameter, we indicate the default value used, the range in the sensitivity analyses, and the references.

| Parameter | Value retained | Range for SA | Reference |
| --- | --- | --- | --- |
| Omicron generation time | Gamma(shape=1.84, rate=0.53) | See Appendix S4 | UKSHA [2022] |
| Omicron VOC decrease in virulence | 0.33 | Not included | Nyberg et alii [2022] |
| Intrinsic transmissibility | Fitted on data |  |  |
| Transmission reduction | 0.5 | [0.45 – 0.55] | Bosetti et alii [2022] |

### Vaccinal efficacy and immunity waning

In contrast to earlier models, we could calibrate immunity waning using epidemiological data (Figure 1B).

The UKHSA reports provide time series for vaccine efficacy over time for the different variants UKHSA [2022]. They also provide estimates of vaccine effectiveness against symptomatic disease, which we used as protection against infection in our model (we assumed no difference between age groups for this parameter). We also used their vaccine effectiveness time series against hospitalisation to parameterize variations in protection against severe cases in our model. This waning was assumed to differ between individuals younger or older than 60 y.o. The implementation was done by assuming a linear model for the decrease in immune protection, as detailed in Appendix S4.

Note also that although we had access to vaccine efficacy for both two-doses vaccinated and boosted individuals, we could not find similar data for recovered individuals. For simplicity purposes, we assumed that recovered individuals had the same level of immunity as two-doses vaccinated individuals. This hypothesis may overestimate the level of protection of natural infections against infection by another variant. We moreover assumed that previously hospitalised individuals had higher protection than individuals that developed mild symptoms, as it is reported they have higher antibody titers [Servellita et alii, 2022].

### Past vaccine rollout and prospective future vaccination campaigns

As in many high-income countries, in France, the first vaccination campaign started in early January 2021. In our model, the vaccine rollout was fitted by age groups accordingly to real data (see Reyné et alii [2022] for details). The youngest individuals (0-10 years old) were not eligible for vaccination in this model.

For this initial vaccination campaign, we assumed moreover an automatic third (booster) dose six months after the second one, meaning that every individual leave the *V*_*a*_ compartment to enter the boosted compartment (*B*_*a*_).

The future (and prospective) vaccine campaigns — presented hereafter — concern only individuals in the booster compartment (*B*_*a*_). We assumed perfect population compliance to future vaccination from the individuals already vaccinated, meaning that everyone eligible receives his booster vaccine dose. This assumption is likely to be optimistic but it is also counterbalanced by the fact that we also assumed unvaccinated individuals will remain as such.

We investigated three different scenarios:

- In **Scenario A**, boosted individuals are not vaccinated again.
- **Scenario B** consists in implementing annual vaccination campaigns before winter (in September and October) but only for individuals above 60 years old.
- We extend the yearly vaccination campaign to all the population in **Scenario C**.

## 3 Results

Scenario A leads to a high level of daily hospital admissions (Figure 2A) with yearly oscillations attributable to seasonality.

**Figure 2:**
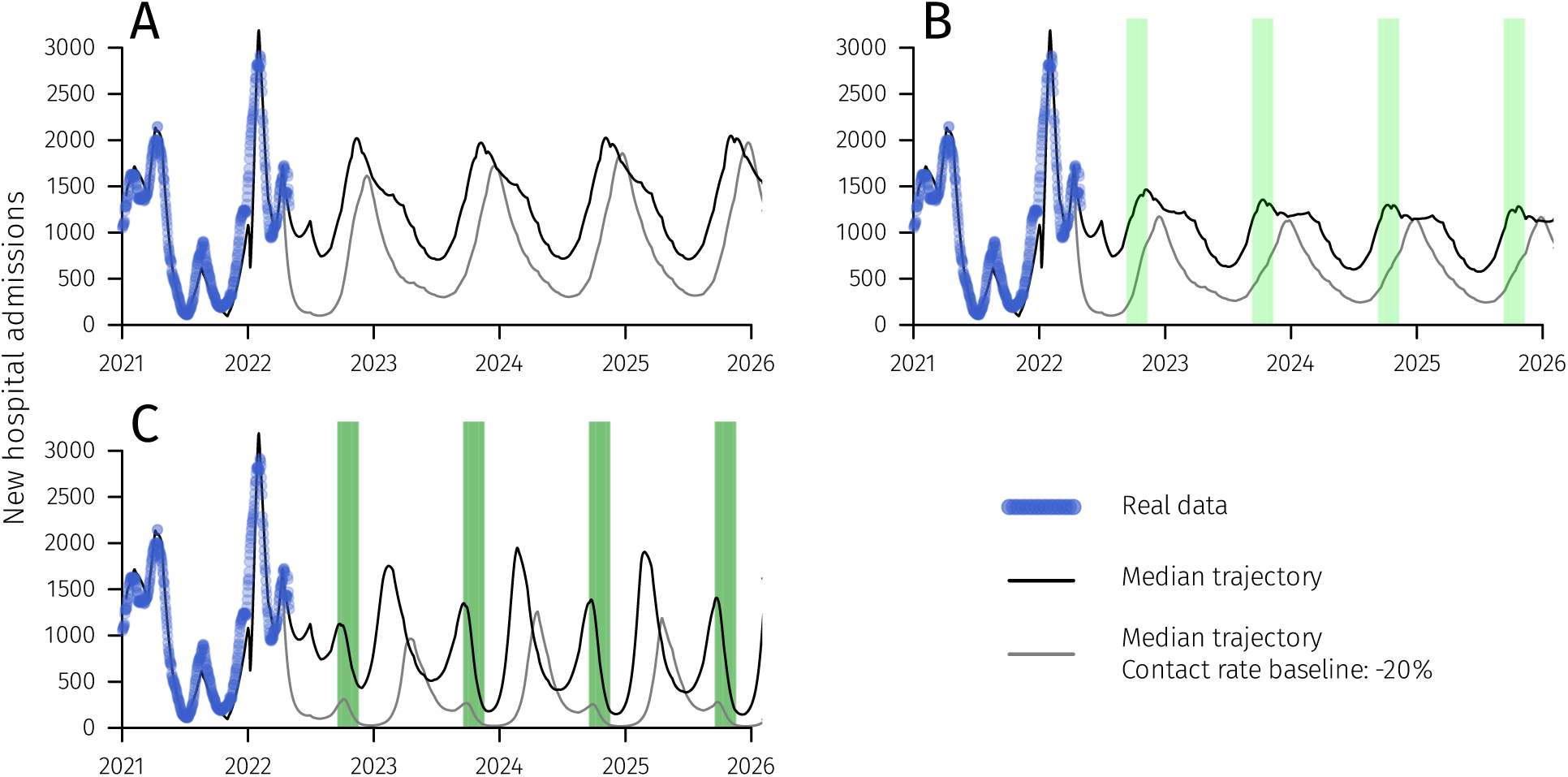
Median number of COVID-19 hospital admissions in four vaccination campaign scenarios. Blue dots corresponds to real data and green shaded areas to vaccination campaigns periods. The four panels correspond to scenarios without additional booster **(A)**, with an annual vaccination campaign in September-October for every individuals above 60 y.o. **(B)**, an annual vaccination campaign in September-October for the whole population **(C)**. See Supplementary Figures S1 to S3 for the 95% confidence intervals and the interquartile range.

Scenario B improves the overall situation but the median number of hospital admissions always remains above 500 per day (Figure 2B).

Vaccinating everyone once a year (Scenario C) further lowers the number of daily hospital admissions and it also yields an epidemic wave in the early spring (Figure 2C).

Scenario C exhibits the most pronounced epidemic waves, which appear to occur twice a year. This can be explained by the fact that vaccinating everyone at the same time implies that immunity wane for everyone at the same time. This generates two periodicities, one related to seasonality, and the second related to vaccination campaigns.

Figure 3 shows the annual total number of hospital admissions for each scenario. As expected, the more people are vaccinated, the more the daily hospital admissions lower. However, vaccination alone just seems to contain what was reached with stringent NPIs in 2021 in France (curfews, lockdown, health pass).

**Figure 3:**
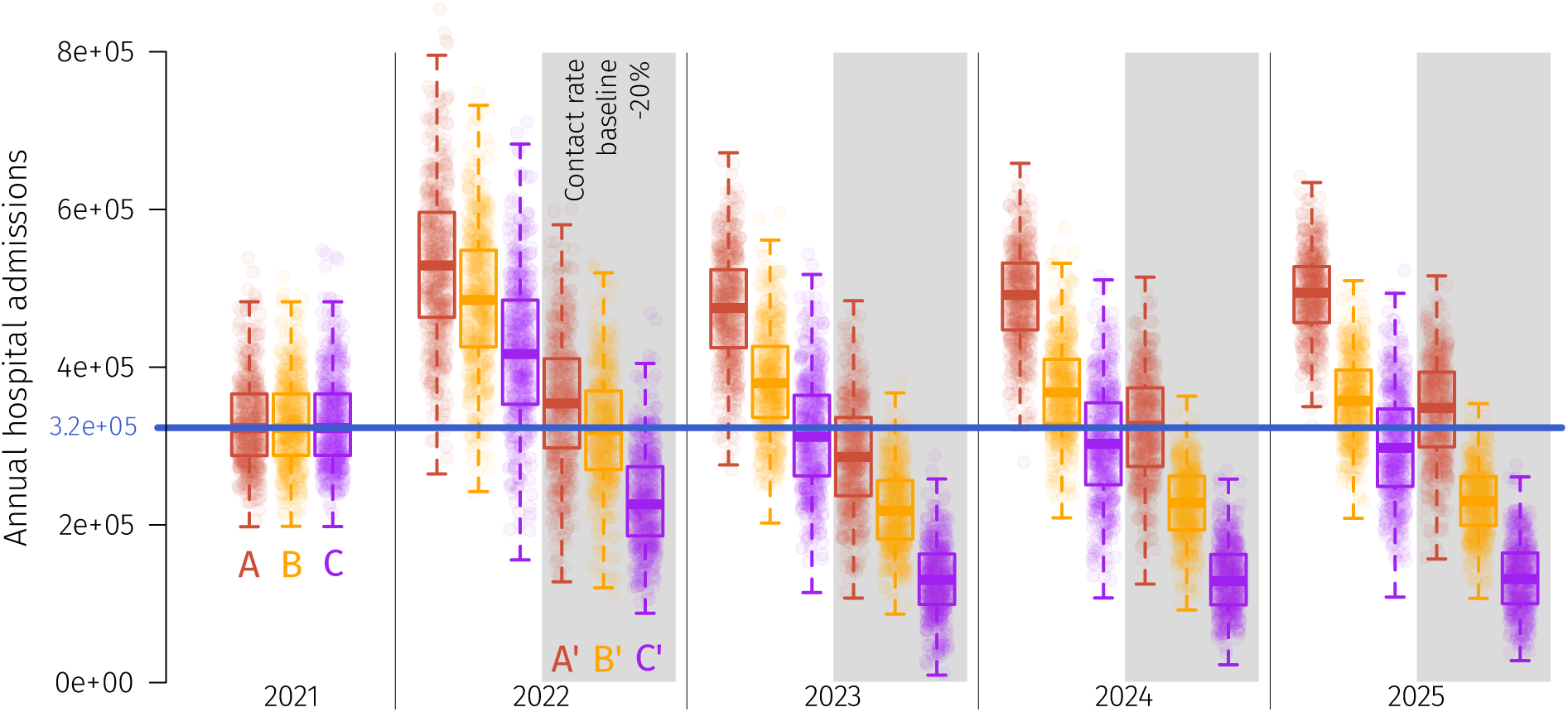
Comparison of the total number of hospital admissions per year for four vaccination scenarios. Blue line corresponds to real data for 2021. The different type of boxplots correspond to the different scenarios. Scenarios **A’, B’** and **C’**, in the gray shaded area, corresponds to Scenarios A, B and C where the baseline transmission rate was decreased by 20%. Complete dynamics for those alternate scenarios are shown in Supplementary Figures S4-S5.

Note that, as shown also in Supplementary Figures S1 to S3, simulations yield large 95% confidence intervals (CI) for the total number of hospital admissions.

Finally, we explored the impact of a 20% reduction in the baseline transmission rate for the three scenarios (Supplementary Figures S4-S5). On its own, such a reduction in transmission rates can have a stronger impact than yearly vaccination campaigns for the whole population (scenario A’ in Figure 2E). Combining yearly vaccination and a decrease in transmission rate provides the strongest decrease in the total number of yearly hospital admissions.

## 4 Discussion

COVID-19 management now faces two major challenges: first the post-infection and post-vaccination immune waning and, second, the emergence risk of new VOCs. These two factors amplify the level of uncertainty and unpredictability inherent to long-term predictions. However, more than two years after the first outbreak, five, six, or seven epidemic waves later depending on the countries and six million deaths world-wide, we believe it necessary to better apprehend long-term dynamics. It would provide valuable insights for health policy decision-making in comparing different strategies.

Although the nature of future VOCs is largely unpredictable, mathematical modelling can incorporate the heterogeneity in immune protection observed in vaccinated populations by using alternate formalism that accounts for the time spent in each compartment and thus implements a realistic loss of immunity over time. We apply this approach to the case of France, which resembles that of many high-income countries, by leveraging empirical Omicron-based data on immune protection.

Despite a huge uncertainty inherent to this approach, we obtain long-term qualitative COVID-19 hospital admissions trajectories with noticeable differences between scenarios. In most of our scenarios, we find that immune waning may cause yearly epidemic hospitalisation peaks comparable to the largest one seen in 2021. Although, as expected, vaccination decreases the total number of hospital admissions, our simulations suggest they are insufficient to suppress the epidemic and the public health burden by itself.

By comparing scenarios with different levels of baseline NPIs, we show that all other things being equal, policies decreasing the transmission rate by 20% can lead to a decrease in hospitalisations comparable to yearly vaccination of the whole population (Figure 3). In comparison, mask-wearing is reported to have a similar order of magnitude, with a reported reproduction number reduction of ~19% [Leech et alii, 2022]. We also show an effect in that the combined implementation of vaccination and NPIs have the greatest impact on reducing the hospital burden (Figure 3).

These results should be taken with caution and regarded as qualitatively prospective due to the numerous sources of uncertainty (Appendix S1). Moreover, this study presents limitations induced by the strong biological assumptions summed up in Box 1. For these reasons, comparisons should be restricted to our different scenarios that share the same core assumptions.

The sensitivity analyses (Figures S6-S8) highlight the main sources of uncertainty which correspond to factors that are difficult to predict. As discussed by Reyné et alii [2022], the time-varying contact matrix yields a huge uncertainty in the model’s outputs but is one of the most difficult model components to parameterize —as it depends on government policies, age-specific spontaneous behavioural changes or calendar events such as school holidays. Seasonality also impacts strongly the results and its precise effect is still under investigation. Regarding virus-related model parameters, the reduction in contagiousness due to immunity, which is difficult to estimate [Bosetti et alii, 2022; Prunas et alii, 2022], does contribute to a large proportion of the variance in the model output (which is the daily number of newly hospitalised individuals).

Some factors are not included in the model but could affect the dynamics. For instance, this study does not address virus evolution on a long-term scale although five VOCs have already spread in France in 2021. It also neglects the potential hospitalisations attributable to patients with long-COVID.

Finally, we assumed that the intensity of non-pharmaceutical interventions will remain identical to that enforced in early spring 2022. This neglects any changes in government policies, some of which would probably be necessary to avoid hospital saturation for some parameter sets.

Overall, in spite of its limitations, this work is, to our knowledge, one of the first to build on empiric estimates of immune protection waning to provide us with long-term perspectives. Its results underline the potential interest of combining vaccination with other types of interventions, especially NPIs such as improving indoor air quality or mask-wearing, to minimise the COVID-19 burden on hospitals in high-income countries. Future work could help to identify the optimal schedule for COVID-19 vaccination campaigns, which will require narrowing many unknowns regarding the biology and spread of the virus.

### Box 1

**Main biological assumptions, simplifications, and limitations of the model**

As with any mathematical model, the one developed in this study makes several assumptions regarding the biological processes at work to be able to interpret the results in a meaningful manner. The main limitations are hereafter listed:

- The **life histories of immunised hosts are limited to few patterns**. Indeed, we assume that all individuals in a given compartment have the same protection against the infection and the disease but, in reality, there is heterogeneity driven by the number of vaccine doses, infections, and their order [Reynolds et alii, 2022]. However, thanks to the non-Markovian structure of the model, we do capture temporal heterogeneity (i.e. protection waning).
- We assume **a perfect compliance of population to vaccination**, thus ignoring a potential ‘fatigue’ in the population [Bodas et alii, 2022; Di Domenico et alii, 2021]. We also assume an **efficient vaccine rollout**, even though logistical issues might emerge.
- There is **no virus evolution** in the model. However, on such a long-term scale, new phenotypic variants will likely emerge.
- There is **no spatial structure**, which could artificially increase the magnitude of the yearly epidemic waves [Thomine et alii, 2021].

## Data Availability

The scripts will be available upon publication.

## Acknowledgements

The authors acknowledge the ISO 9001 certified IRD i-Trop HPC (South Green Platform) at IRD Montpellier for providing HPC resources that have contributed to the research results reported within this paper. We thank all the ETE modelling team for discussions and apologize to Baptiste Elie for that reckless `rm -r *`.

## Statements and Declarations

### Authors’ Contributions

All authors contributed to the study conception and design, interpreted the results and wrote the manuscript. Analysis were performed by BR. All authors read and approved the final manuscript.

### Data availability

Data and scripts will be available upon publication in the following git repository https://gitlab.in2p3.fr/ete/long_term_covid.

## S1 Supplementary figures

**Figure S1:**
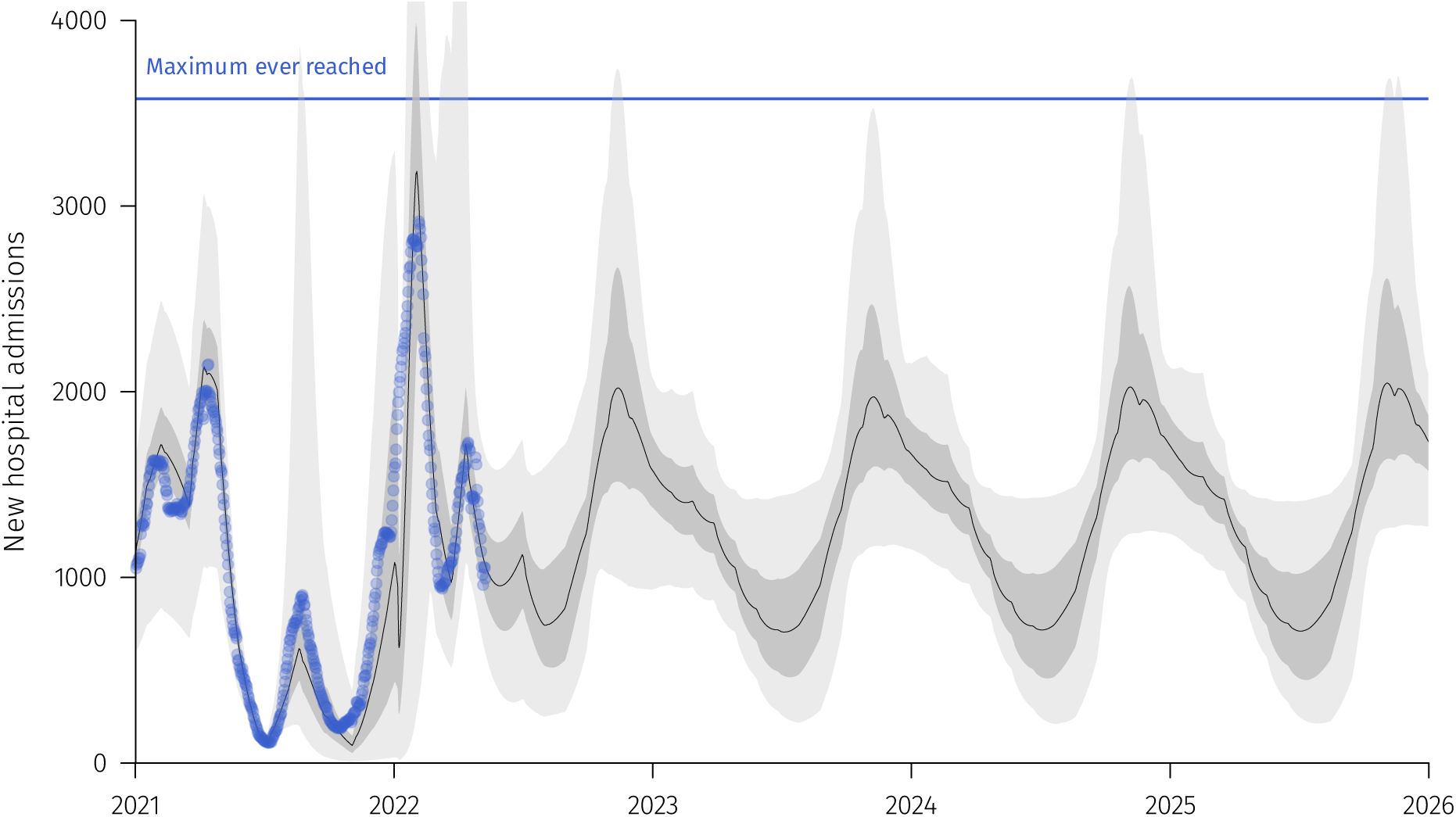
Detailed output of Scenario A (no additional vaccination). Blue dots corresponds to real data. The black line correspond to the median trajectory. Lighter shaded area correspond to the 95% confidence interval while the darker area correspond to the interquartile range. The horizontal blue line indicates the highest national incidence in hospital admissions reached during the first COVID-19 wave in France.

**Figure S2:**
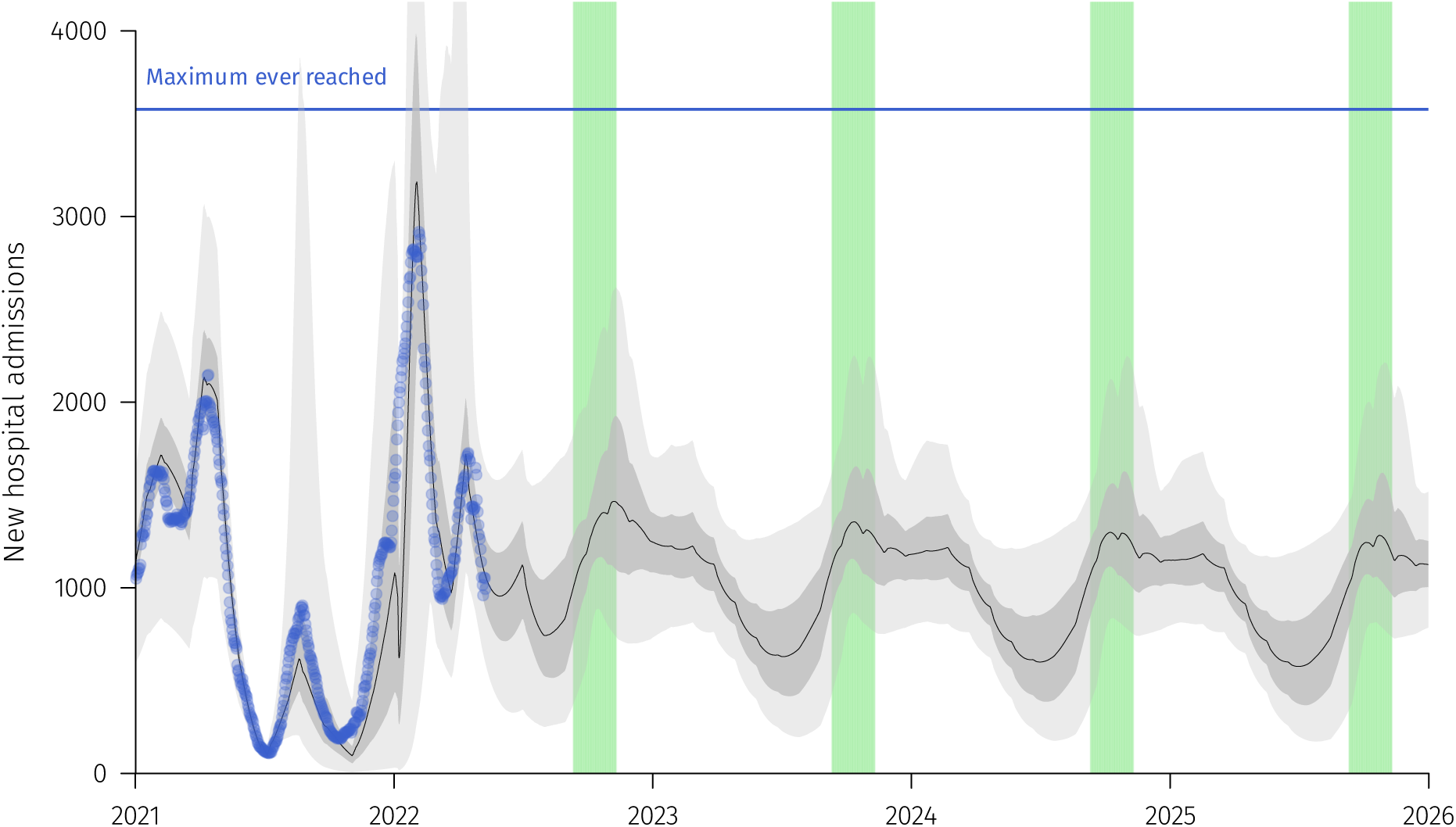
Detailed output of Scenario B (yearly vaccination of individuals of more than 60 years old). Green shaded areas correspond to vaccination campaigns periods for individuals above 60 y.o. See Figure S1 for additional details.

**Figure S3:**
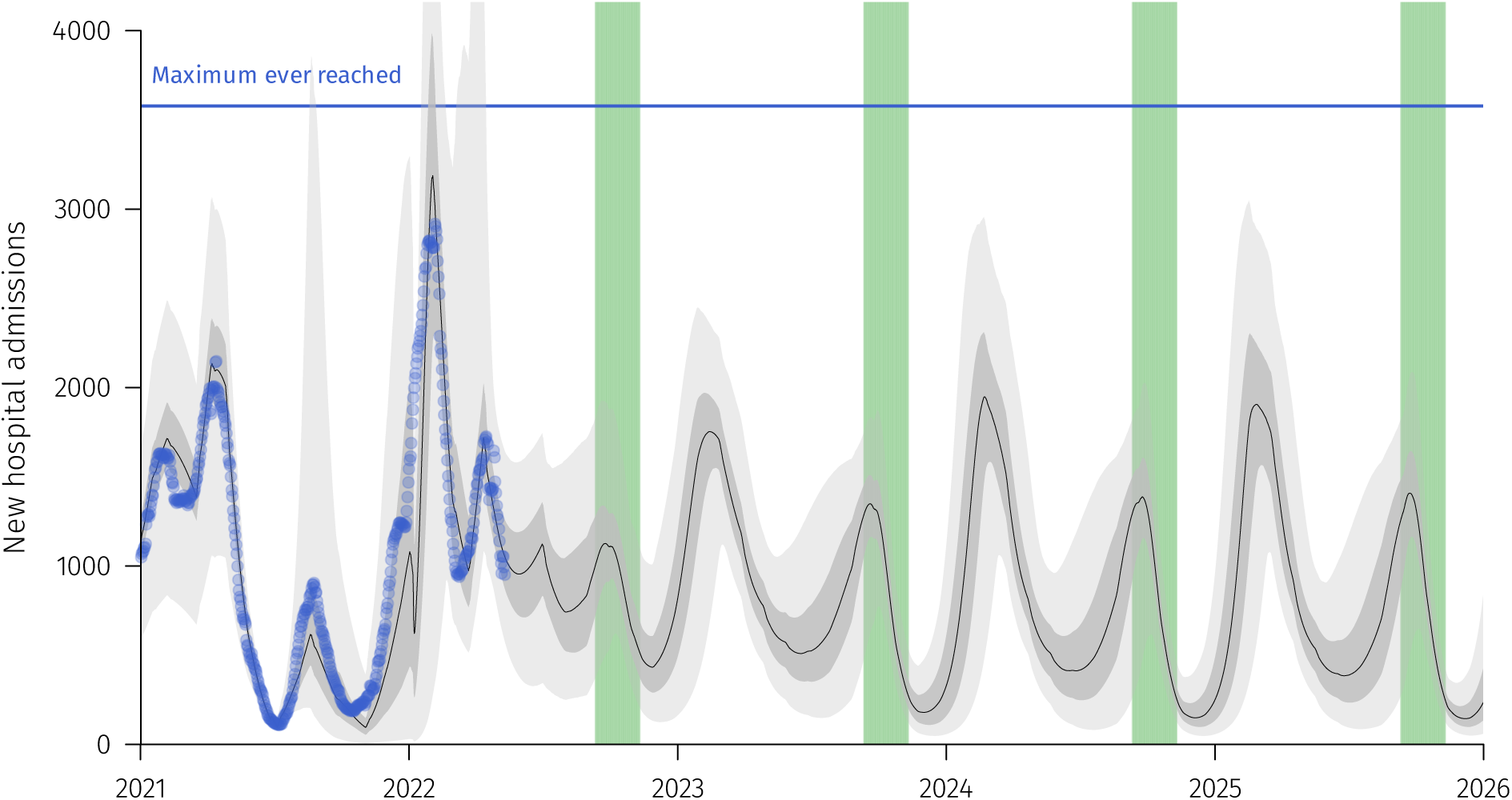
Detailed output of Scenario C (yearly vaccination of all the population). See Figure S2 for additional details.

**Figure S4:**
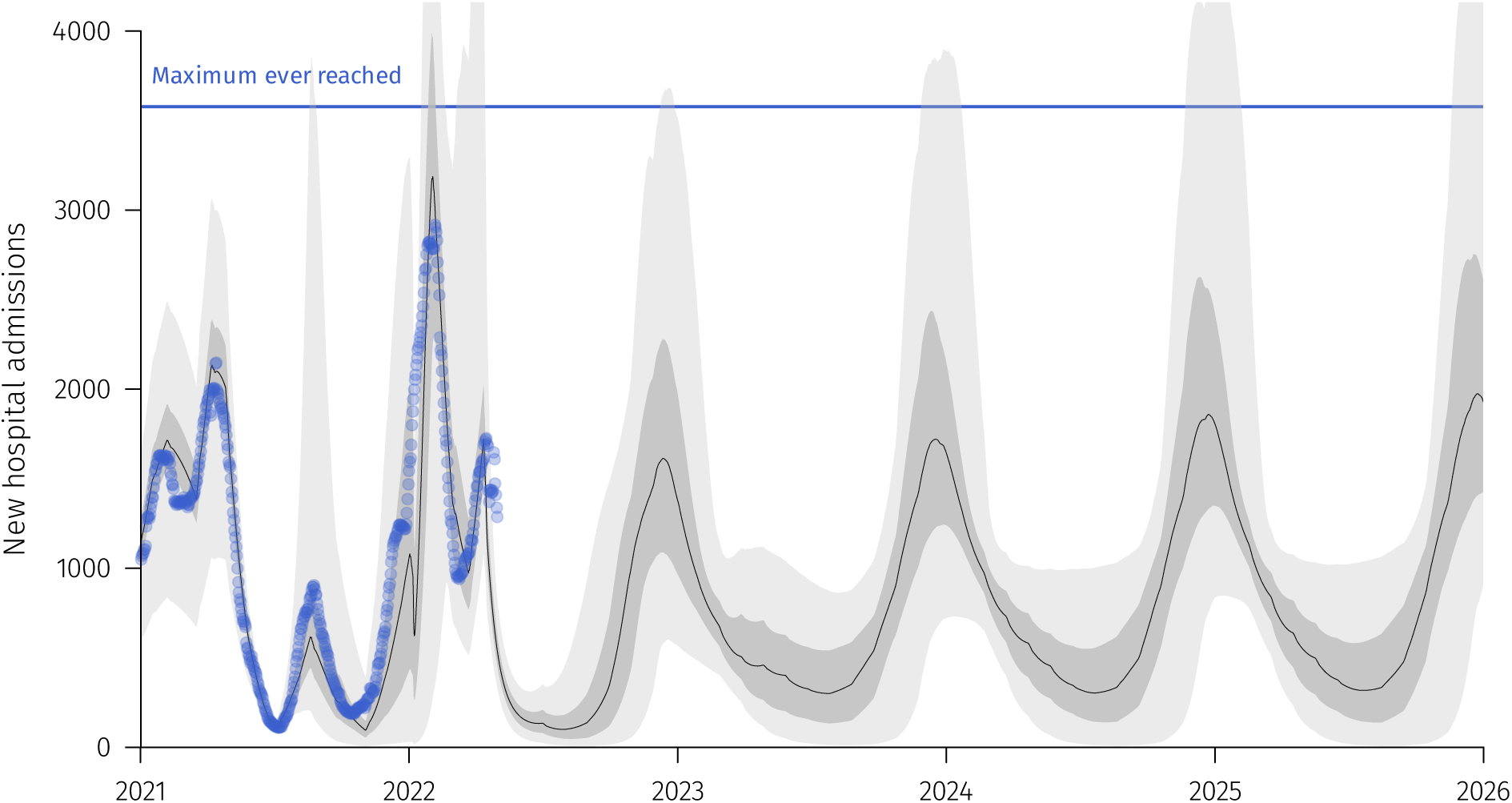
Detailed output of Scenario A (yearly vaccination of all the population) with a 20% reduction of the transmission rate. See Figure S2 for additional details.

**Figure S5:**
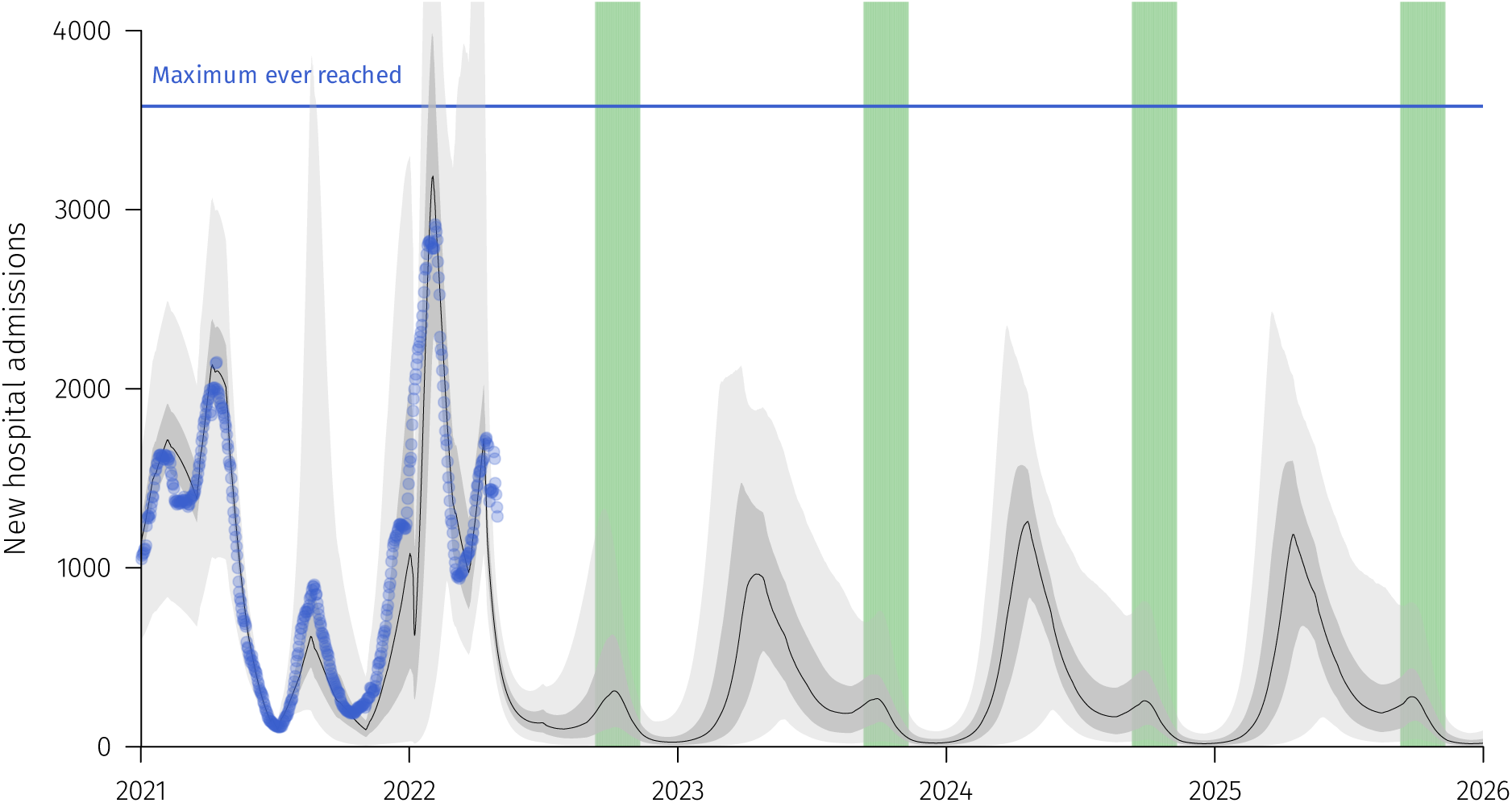
Detailed output of Scenario C (yearly vaccination of all the population) with a 20% reduction of the transmission rate. See Figure S2 for additional details.

**Figure S6:**
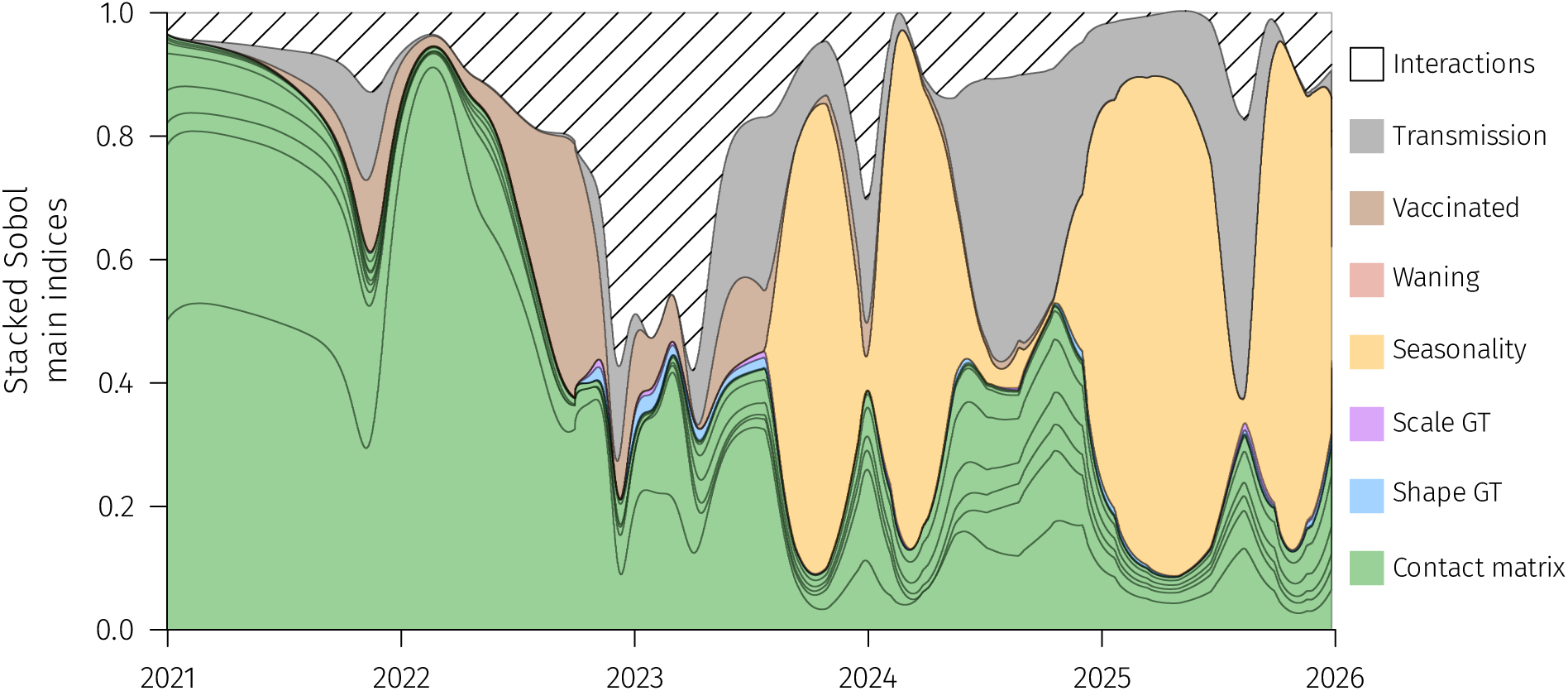
Global sensitivity analysis for Scenario A. The graphs shows the origin of the variance captured by each model parameter using Sobol indices (see Reyné et alii, 2022 for methodological details). A large part of the variations originates from the unknowns in the contact matrix between ages, as in Reyné et alii, 2022. The magnitude of seasonality also matters for the long-term trends, as well as the transmission rate (i.e. the intensity of the NPIs).

**Figure S7:**
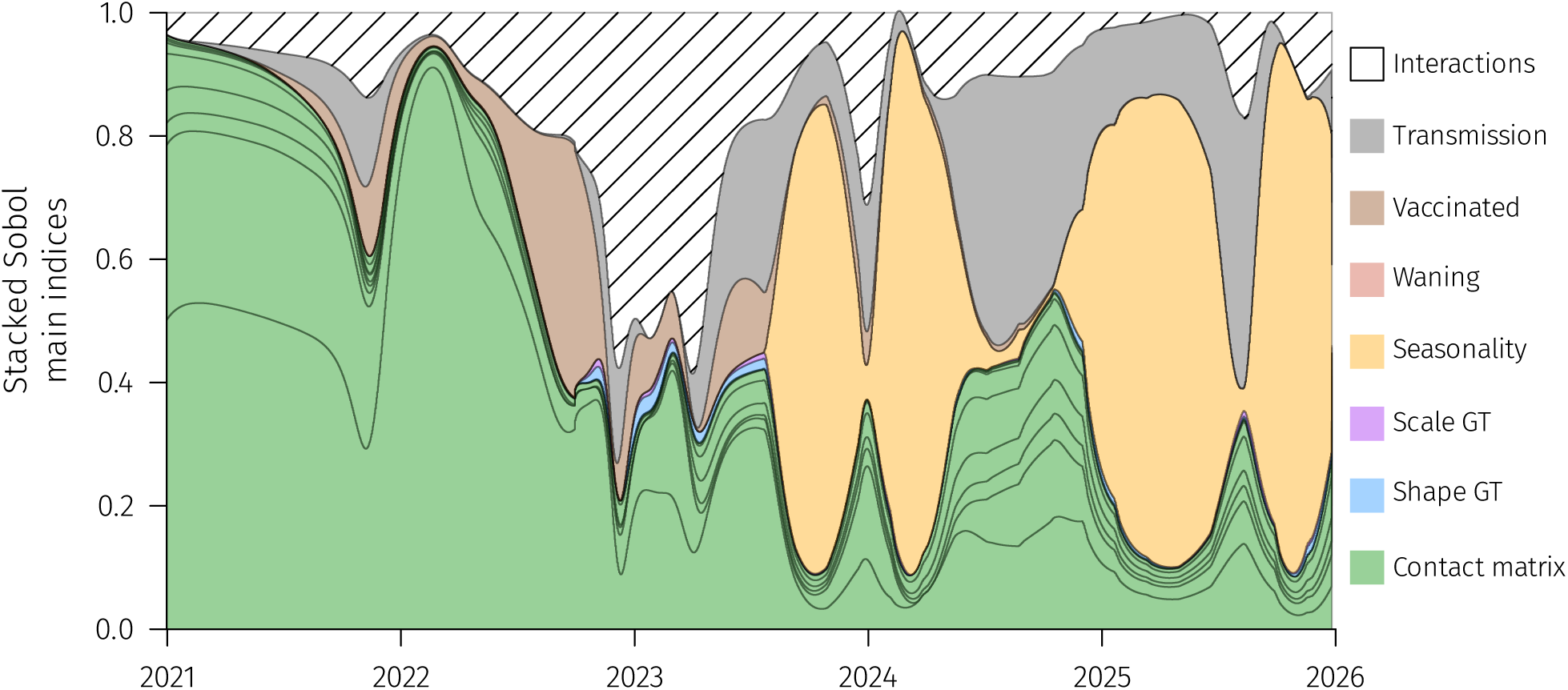
Global sensitivity analysis for Scenario B. See S6 for details.

**Figure S8:**
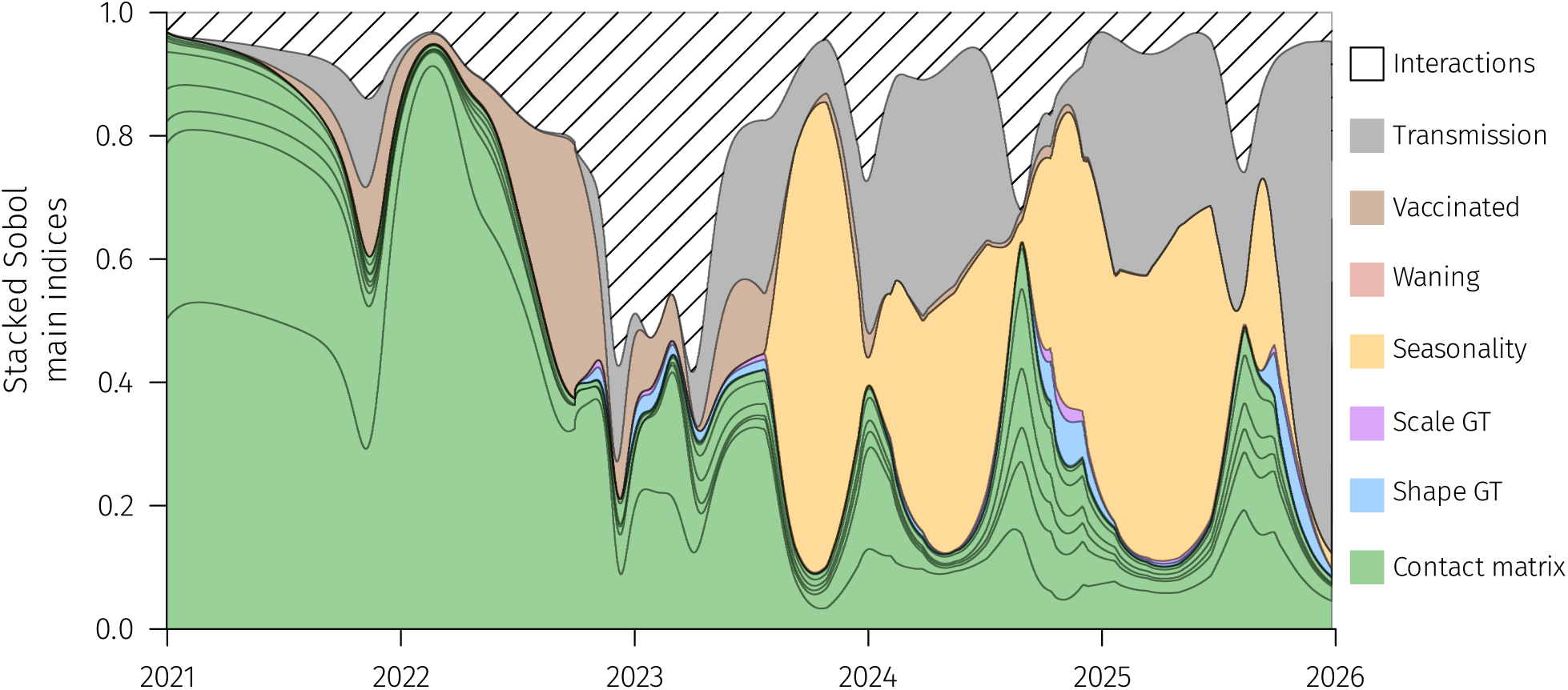
Global sensitivity analysis for Scenario C. See S6 for details.

## S2 Model equations

The model partial differential equations system is given by:

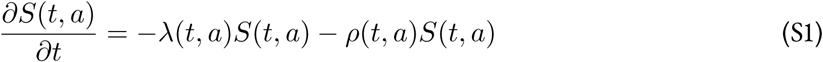

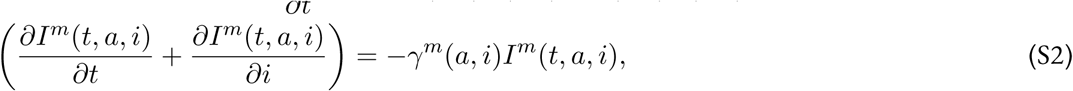

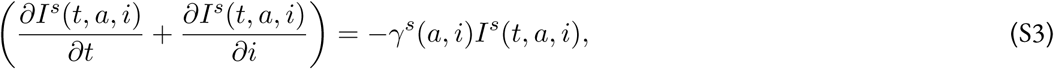

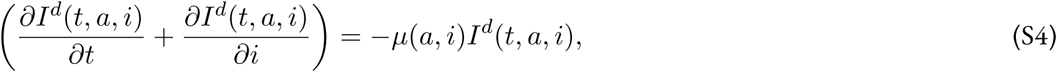

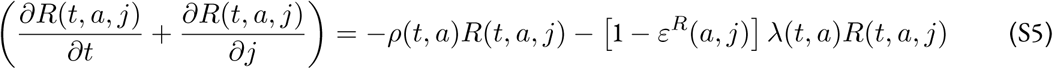

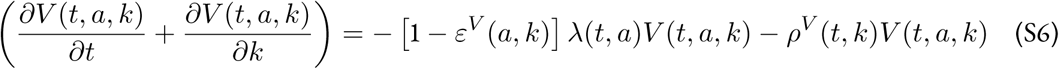

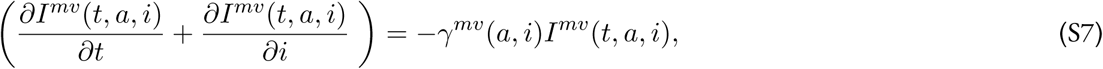

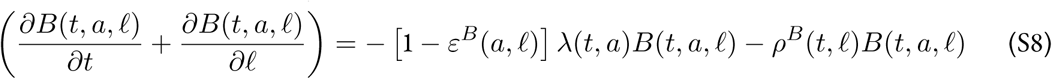

with

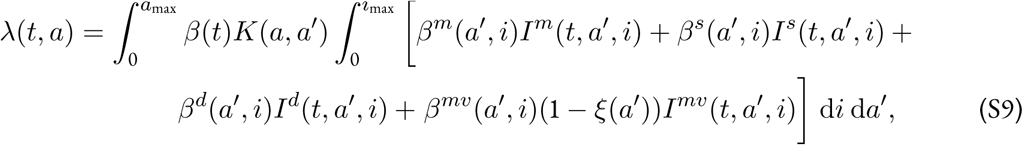

for any (*t, a, i, j, k*) ∈ ℝ^+^ × [0, *a*_max_] × [0, *i*_max_] × [0, *j*_max_] × [0, *k*_max_].

The parameter notations are the following:

- *ρ* is the initial vaccination rate,
- *ρ*^*V*^ is the first booster dose vaccination campaign,
- *ρ*^*B*^ is the re-vaccination rate of prospective vaccination campaigns,
- *γ*^*m,s*^ the recovery rates for respectively mildly and severely infected individuals,
- *µ* is the death rate,
- *ε*^*R,V,B*^ the immunity-induced reduction of risk of infection for individuals respectively in the *R, V* and *B* compartments,
- *β*(*t*) transmission rate, accounting for NPIs policies,
- *K*(*a, a*^′^) the contact matrix coefficient between age groups *a* and *a*^′^,
- *β*^*m,s,d,mv*^ the generation time distributions, and
- *ξ* the immunity-induced reduction in transmission (for ‘breakthrough’ infections).

The previous system is coupled with the following boundary conditions:

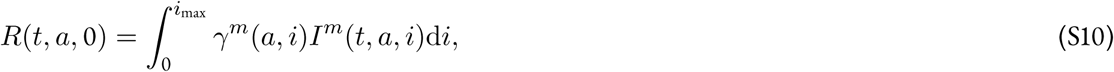

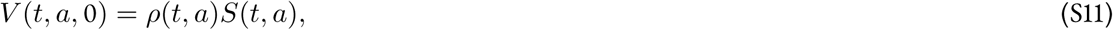

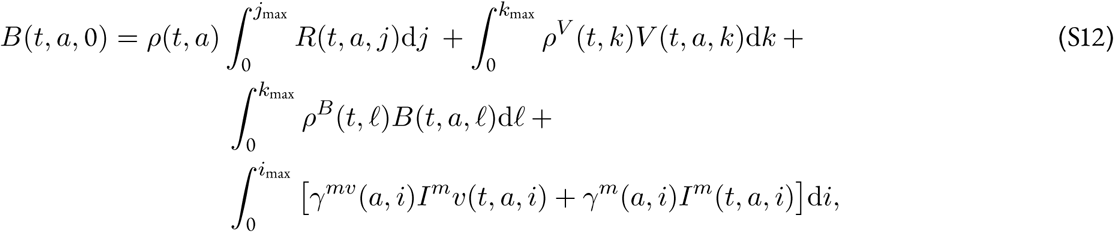

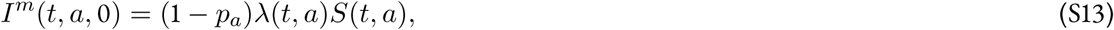

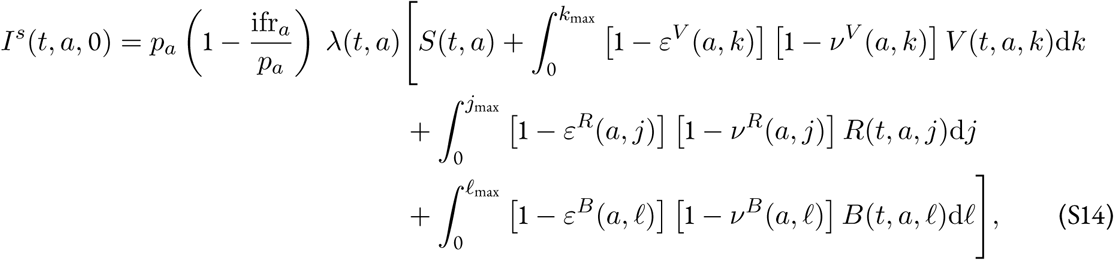

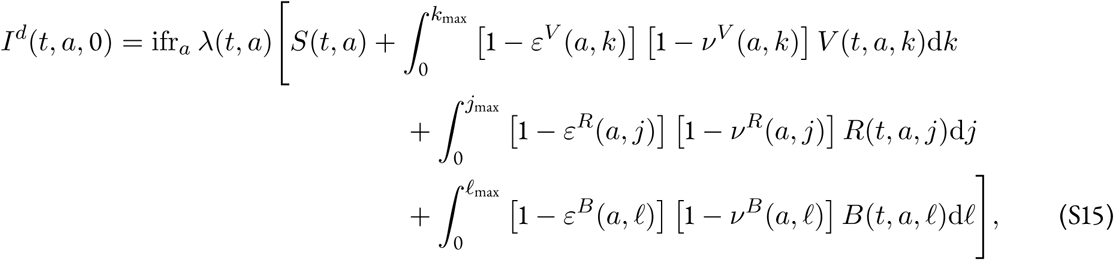

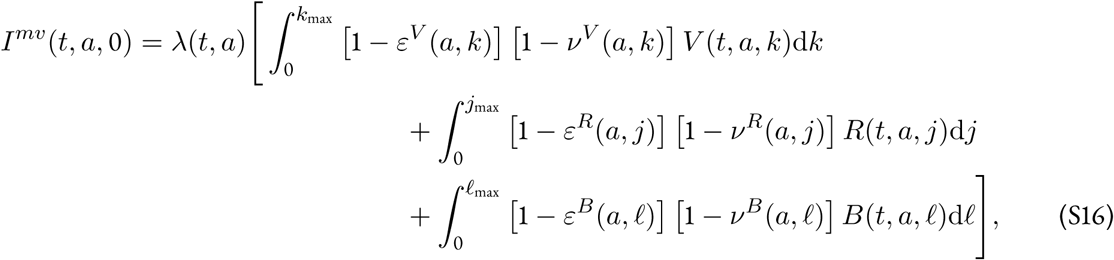

with

- *p*_*a*_ is the probability of developing a severe form,
- ifr_*a*_ is the infection fatality rate, and
- *ν*^*V,R,B*^ is the immunity-induced reduction of virulence for individuals respectively in the *R, V*, and *B* compartments.

## S3 Model parameters

For the other model parameters, they are identical to Reyné et alii [2022]. We sum up here the different parameters used in the model. However, we encourage readers to check Reyné et alii [2022] for the details regarding the contact matrixes, that will not be described in detail here. Long story short, we use 39 different France-specific contact matrixes corresponding to diverse NPIs implementation thath happened in France.

For the others parameters, you can refer to the Table S1.

**Table S1:** Model parameters. For each parameter, we indicate the default value used and the references used.

| Parameter | Value retained | Range for SA | Reference |
| --- | --- | --- | --- |
| Alpha/Delta generation time | Weibull(2.83, 5.67) | Not included | Ferretti et alii [2020] |
| Proportion of severe cases ($p_a$), IFR ($\text{ifr}_a$) | 0.0113 (mean), 0.0022 (mean) | Not included | Verity et alii [2020] |
| $\alpha$ VOC increase in virulence | 1.65 | Not included | Challen et alii [2021]; Davies et alii [2021] |
| $\delta$ VOC increase in virulence | 2.08 | Not included | Fisman and Tuite [2021] |
| Seasonality | 0.1 | [0 – 0.2] |  |
| Mild recovery rate | Adapted | Not included | Salje et alii [2020]; Reyné et alii [2022] |
| Severe recovery rate | Adapted | Not included | Salje et alii [2020]; Lefrancq et alii [2021]; Reyné et alii [2022] |
| Vaccination rate | Fitted | — | See Reyné et alii [2022] |
| Total number of vaccinated (initial two doses) | $5.21 \cdot 10^7$ | $[5 \cdot 10^7 - 5.42 \cdot 10^7]$ | French Minister of Health website (visited on 2021-10-28) |
| Total reduction in virulence | See Appendix S4 | +/- 5% | UKHSA [2022] |
| Infection immunity | See Appendix S4 |  | UKHSA [2022] |
| Transmission reduction | 0.5 | [0.45 – 0.55] | [Bosetti et alii, 2022] |
| Initial proportion of recovered | 0.149 | Not included | Hozé et alii [2021] |
| Age structure | Real data | — | https://www.insee.fr/fr/statistiques/2381474 |
| Contact matrix | — | See Reyné et alii [2022] | Béraud et alii [2015]; Reyné et alii [2022] |

## S4 Omicron related parameters

Regarding the omicron generation interval, we used the data provided by UKSHA [2022]. In particular, we fitted different Gamma distributions on the non-parametric data available. We tested different parameter combinations for the Gamma distribution to explore a range of generation time distributions that reflect the epidemiology of both BA.1 and BA.2 variants. In Figure S9, we show a subset of these Gamma distributions explored within the sensitivity analysis. The selected baseline is Gamma distribution with a 1.84 shape parameter and a 0.53 rate parameter.

Note also that the intrinsic virulence of Omicron is assumed to be divided by 3 (parameter *p*_*a*_ in the model) compared to Delta following UK data. This is mostly qualitative and was not age-differentiated [Nyberg et alii, 2022].

Finally, note that we did not set up an intrinsic basic reproduction number (ℛ_0_) for the Omicron VOC. While it was still possible for the Alpha VOC and arguably for the Delta VOC to have some estimates, the ℛ_0_ only refers to a completely naive population. Hence, when the transmission advantage of a VOC is partly due to an escape of the already built population immunity, it becomes hazardous to retrieve ‘the true ℛ_0_’ for such a VOC. This is the case for the Omicron VOC. Therefore, we only fit the transmission rate on hospital admissions time series, rendering it difficult to dissociate the temporal reproduction number and the NPIs/changes in the behaviour of the population.

**Figure S9:**
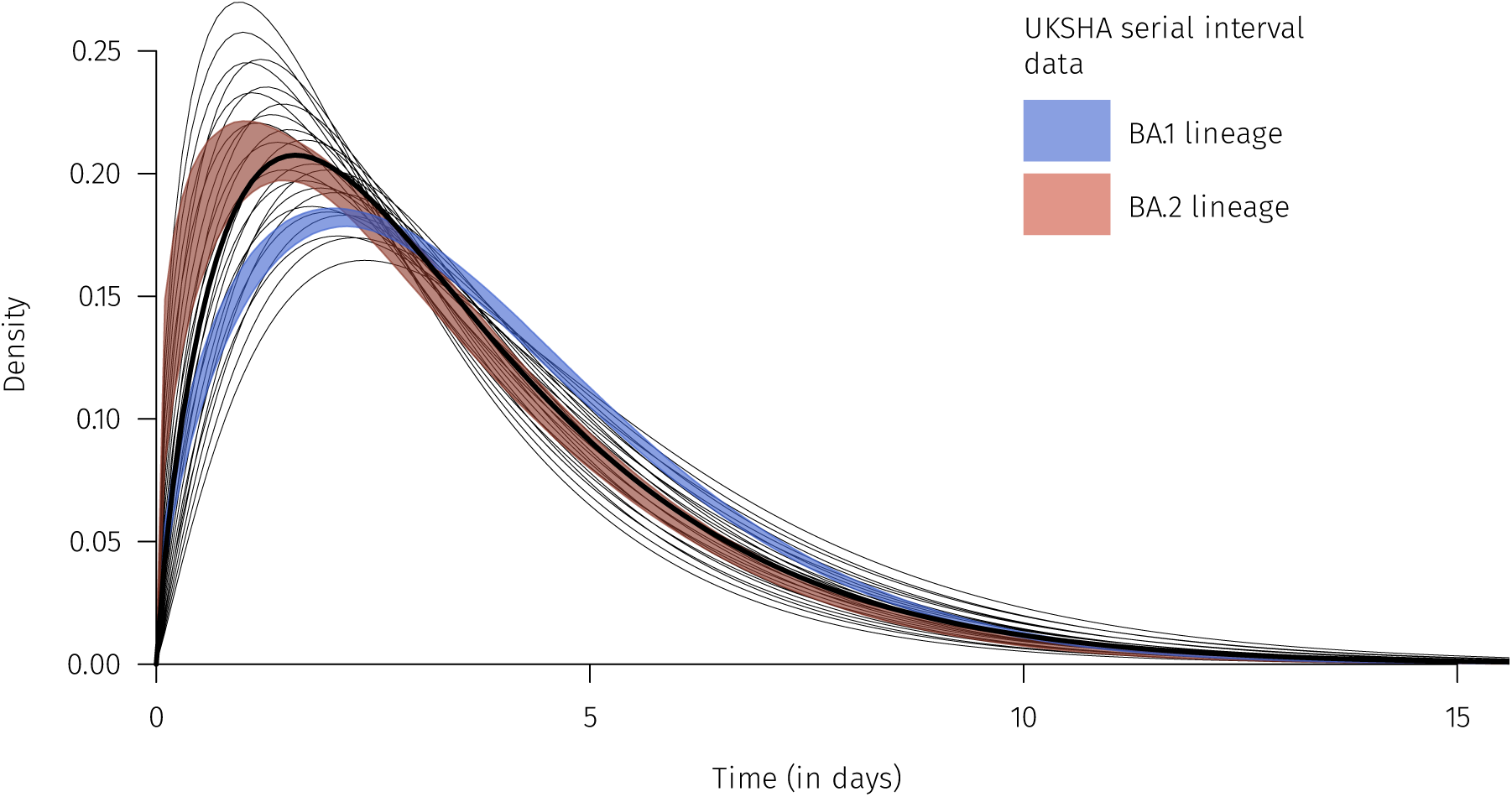
Omicron generation time. Shaded areas data originate from the UKSHA [2022] report. The lines correspond to a subset of the Gamma distributions used in the sensitivity analysis. The thicker line corresponds to the baseline Gamma distribution.

## S5 Vaccine efficacy

The vaccine acts in three different ways in this model.

First, the vaccine protects against infection. (This corresponds to the *ε*^*V/R/B*^ parameters in the model, and accounts for the time since the entry in the compartments.) We used data provided by the UKHSA [2022] report (https://assets.publishing.service.gov.uk/government/uploads/system/uploads/attachment_data/file/1070356/Vaccine-surveillance-report-week-16.pdf). The raw data was not available so we used the online tool WebPlot Digitalizer (https://apps.automeris.io/wpd/) to retrieve the values. For simplicity, we assumed that individuals received the Pfizer/BioNTech (BNT162b2) vaccine, which was the most widespread in France. On each time series (protection against Delta or Omicron, whether the individual received two-dose or a supplementary booster dose), we fitted a linear model (cf. Figure S10).

Second, the vaccine protects also against severe forms if individuals are nonetheless infected. (In the model, the probability of developing a severe case *p*_*a*_ is decreased by (1−*ν*^*V/R/B*^), which also accounts for the entry in the *V/R/B* compartments.) The same methodology is done as previsously, with the difference that UKHSA data show the total protection against virulence, *i*.*e*. [1−*ε*^*V/R/B*^(·)] [1−*ν*^*V/R/B*^(·)]. Another difference is that we distinguish between individuals above and below 60 y.o. as shown in Figure S11. One last thing to note is that we assumed that the last data point for individuals below 60 y.o. that received a booster was the upper limit provided and not the baseline value since the baseline for boosted individuals was below tho one with two-doses only.

In the sensitivity analysis, we included variations in the total protection against virulence (cumulative effect of *ε* and *ν*). The variations correspond to add to the intercept of the linear model a coefficient between -0.05 and +0.05.

Finally, the vaccine reduces the infectiousness of infected individuals. The reduction of transmission (in so-called ‘breakthrough’ infections) was assumed to be 50% for vaccinated individuals, as in other modelling works [Bosetti et alii, 2022]. This reduction in transmission was also applied to recovered people that got reinfected. Note that other studies tend to find different estimates [Prunas et alii, 2022].

**Figure S10:**
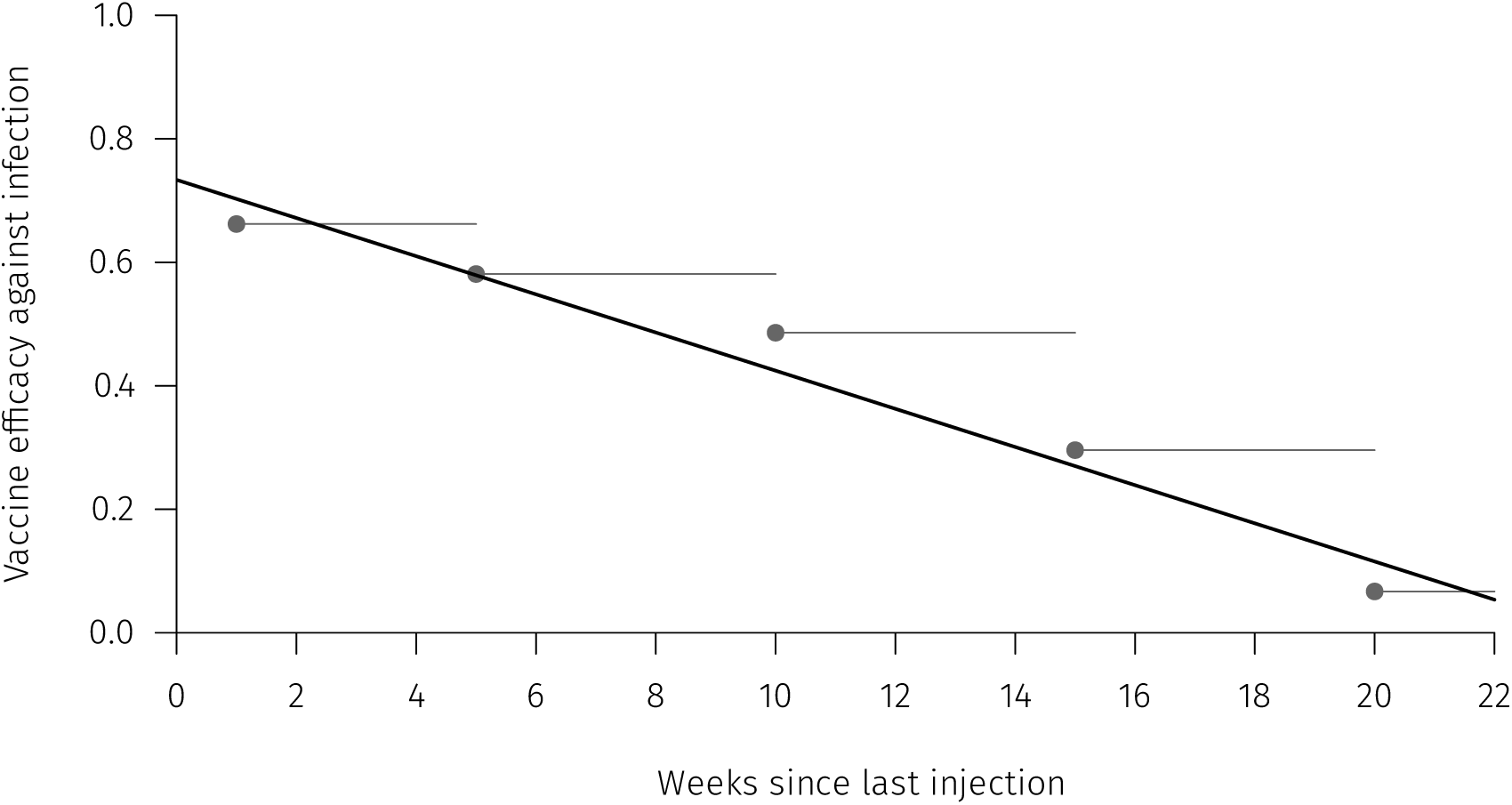
Model implementation of the decrease in immunity following booster vaccination. This is the vaccine efficacy against an Omicron infection after receiving a booster dose compared to fully susceptible individuals. Dots corresponds to real data from UKHSA [2022] for the Pfizer/BioNTech vaccine (BNT162b2) after a booster dose. The line correspond to the baseline of the immunity decrease model implementation.

**Figure S11:**
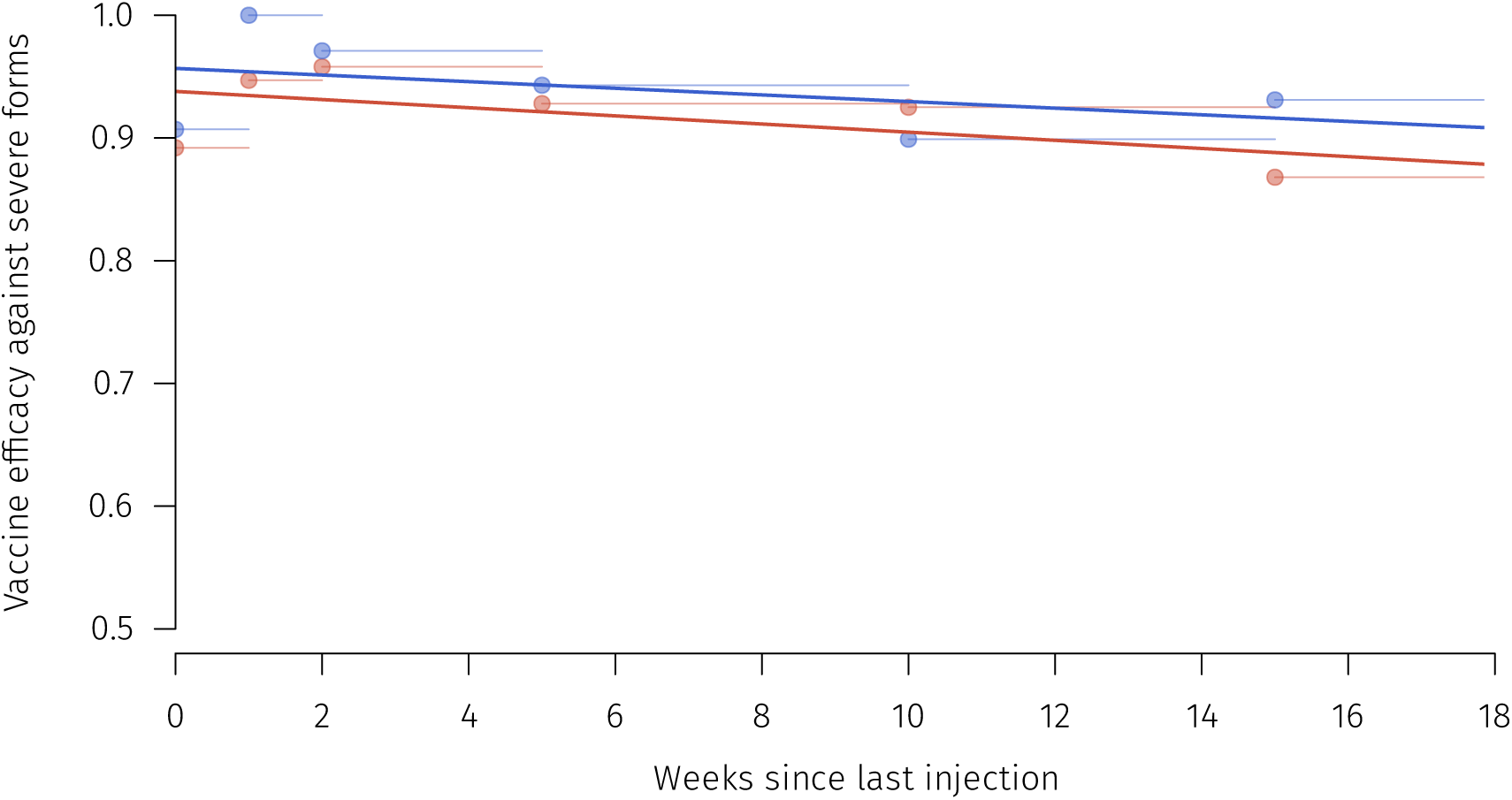
Model implementation of the decrease in immunity following booster vaccination. This is the vaccine efficacy against an Omicron severe case after receiving a booster dose compared to fully susceptible individuals. Dots corresponds to real data from UKHSA [2022] for the Pfizer/BioNTech vaccine (BNT162b2) after a booster dose. Blue color is for individuals below 60 y.o. and red for older people. The lines correspond to the baseline of the immunity decrease model implementation.

